# a-TIGIT mAb belrestotug in combination with anti-PD1 induces an immunocompetent tumor microenvironment (TME)

**DOI:** 10.1101/2025.07.25.25332187

**Authors:** Julia Cuende, Nicolas Rosewick, Valerie Roobrouck, Bart Claes, Paola Tieppo, Marjorie Mercier, Noemie Wald, Anais Vezzu, Julie Preillon, Alizée Canevat, Reece Marillier, Francesco Strozzi, Sophie Brogniet, Marianne Blockmans, Anna-Maria Barbuto, Clarisse Truong, Jeri Kim, Olivier De Henau, Yvonne McGrath, Gregory Driessens, Marion Libouban

## Abstract

Belrestotug, a monoclonal antibody against TIGIT, was evaluated alone and with anti-PD-1 therapy in two clinical trials for advanced solid cancers (NCT04335253, NCT05060432). Belrestotug alone reduced Treg cells and transiently increased proliferating T/NK cells, indicating immune activation in the periphery. Combined with anti-PD-1, it promoted earlier T cell proliferation and further decreased Treg numbers both in blood and tumors, potentially overcoming Treg-related resistance to PD-1 blockade. Spatial analysis demonstrated a reduction of TIGIT+ regulatory T cells within the tumor, along with increased CD8 IFNγ expression and macrophage-associated signatures. These findings indicate that this combination therapy may enhance anti-tumor immune responses by depleting immunosuppressive Tregs and remodeling the tumor microenvironment. Several studies proposed that PD-(L)-1 blockade responses occur mostly in patients with inflamed immunotype, where an enriched immune infiltrate distributes both in tumor nest and stroma regions. Conversely, in excluded immunotypes the immune infiltrate is preferentially localized in the stroma. Our data in late-stage patients, mixed solid tumors, supports that the combination therapy remodels immune cells distribution in the stroma, potentially indicating that inflamed and excluded immunotypes may be predictive biomarkers of response for belrestotug combination with aPD1.

## Introduction

Therapeutic agents that block Programmed Cell Death Protein 1 (PD-1) or its ligand PD-L1 are particularly effective in unleashing anti-tumor CD8 T cell activity, resulting in powerful and durable therapeutic responses against many cancer types. However, only a minority of patients of patients respond to anti-PD-(L)-1s (aPD-(L)-1; aPD-L1 or aPD1) therapies, highlighting the urgent need for combination therapies^1^.TIGIT (T Cell Immunoreceptor With Ig And ITIM Domains) is a member of the poliovirus receptor (PVR)/NECTIN family ^2^ and has emerged as a promising combination partner for a-PD-(L)-1 therapies. In CD8 T cells, TIGIT competes for ligand binding with the costimulatory receptor CD226 also inhibited by PD-1^3^. TIGIT is also expressed by CD4 T cells and natural killer (NK) cells ^4,5^. Among CD4 T cells, most regulatory T-cells (Tregs) express TIGIT constitutively, and its expression identifies a subset with more potent immunosuppressive functions^6^. Comparatively, TIGIT+ Treg cells inhibit more effectively effector T-cell activation than their TIGIT-counterparts^4,7 8^. While PD-1 blockade primarily acts by unleashing the antitumor function of effector T cells, mainly CD8+ T cells ^9^ , different reports suggest it can augment Treg numbers and function in a cell-intrinsic manner, in gastro-esophageal cancer and melanoma patients ^10,11^. Moreover, the Tregs co-expressing TIGIT and PD-1 are more abundant intratumorally than in peripheral blood in NSCLC patients. They are preferentially distributed in the tumor stroma ^12^ further supporting the rational for the combination of a-TIGIT and a-PD-1 inhibitors.

Belrestotug, previously known as EOS-884448, is antagonistic anti-TIGIT monoclonal antibody (mAb) with an Fc enabled region that triggers anti-tumor activity in tumor syngeneic mouse models, by different mechanisms of action. In vitro, belrestotug induces ADCC (Antibody-mediated cell-dependent cytotoxicity) depleting preferentially TIGIT+Tregs^13^. Belrestotug entered clinical development as single agent (NCT04335253) or in combination with standard of care therapies in advanced solid tumors (NCT05060432).

In the clinical studies, we observed belrestotug’s ability to induce immune activation in patients with late-stage solid tumors. Belrestotug induces proliferation of T/NK cells in peripheral blood of patients treated with monotherapy as well as those treated in combination with aPD-1. Belrestotug treatment also reduces peripheral exhausted TIGIT+ CD8 T cells and TIGIT+ Treg, consistent with its ADCC depleting mode of action. Analysis of patient biopsy samples, using tissue spatial technology, showed decreased levels of intratumoral TIGIT+ Tregs within the stroma, especially when these Tregs are located near CD8 T cells, and that such an effect correlated with clinical benefit. To corroborate the spatial relationship between Tregs and CD8 T cells with a method more suitable for clinical application, we took advantage of conventional immunohistochemistry (IHC) method by immunotyping the tumor samples using CD8 and pan-CK staining to locate CD8 cells and discriminate tumor nest from the stromal tissue. We identified both inflamed and excluded immunotypes as potential predictive biomarkers of the combination of belrestotug and a-PD-1, while it is widely accepted that mostly inflamed immunotype is predictive of benefiting from a-PD1 monotherapy. The IO combination also upregulated CD8 IFNγ and TAM (tumor associated macrophages) signatures suggesting a remodeling of the tumor microenvironment. In the CD8 T cell compartment, the dual TIGIT and PD-1 blockade upregulated several differentiated CD8 T cell compartments, as opposed to the reported effect of a-PD-1 monotherapy which markedly elevates one differentiated CD8 T cell subset (the CD8 stress response) ^14^. These observations in patients with late-stage cancers are consistent with a-TIGIT treatment in i*n vivo* mouse models, where we observed elevated intratumoral effector-like subset and a reduction in the exhausted subset.

## Results

### Belrestotug treatment induces changes in immune cell subsets in the peripheral blood of patients

To understand the effect of belrestotug treatment on the different immune cell subsets in our patients, we collected fresh whole blood (WB) before treatment and at different timepoints after treatment with either belrestotug alone or in combination with a-PD-1 (pembrolizumab or dostarlimab). Belrestotug monotherapy elicited a marked increase of 70% in proliferating memory CD8 T cells and a 100% increase in proliferating NK cells relative to baseline. T and NK cells changes were transient and peaked between 1 and 2 weeks after the start of the treatment. Upregulation of Ki67 in CD8 T cells has been reported early after initiation of ICI treatment in melanoma and NSCLC patients, while the effect on NK activation seems linked to TIGIT blockade^15,16,17^. In contrast, the peripheral Treg cell compartment was significantly altered by a 50% reduction compared to baseline and sustained in time (Fig. 1A). Comparatively combination therapy with anti-PD1 advanced T cell proliferation to an earlier stage after initiation of the treatment. Treg depletion was comparable to a-TIGIT monotherapy. These populations were further analyzed using a more granular gating scheme that investigated TIGIT receptor expression. Combination therapy downmodulated TIGIT+ CD8 T and Treg cells in the periphery, though only the total Treg population decreased as opposed to total CD8 T cell population. (Fig. 1AB) TIGIT expression progressively increases during the process of CD8 T cell exhaustion, as illustrated in Fig. 1C, which presents an analysis based on publicly available data from a comprehensive characterization of CD8 T cells in treatment-naïve NSCLC patients^18^. The CD8 T cell exhaustion process has been defined as gradual, and involving several stages^19^ , and arising from two different progenitor subsets. The progenitor subset co-expressing TIGIT and PD-1 is committed to becoming dysfunctional ^20^. Since belrestotug treatment reduces peripheral TIGIT+CD8 T cells, we analyzed the transcriptomic profile of CD8 T cells based on TIGIT expression. We processed PBMCs from 6 treatment-naïve cancer patients, and after a CD8-enrichment step, cells were stained with two different a-TIGIT antibodies, belrestotug labelled and an a-TIGIT antibody (clone 32959) binding to a non-competitive epitope. This staining strategy allows to discriminate between a TIGIT high fraction, targeted by belrestotug treatment and detected in belrestotug treated patients, and a TIGIT low fraction, unaffected by belrestotug therapy (Fig. 1D). Sorted cells were submitted to transcriptomics analysis by Nanostring. Signature scores revealed that exhausted T cells are enriched in the TIGIT high CD8 T cell population, whereas effector T cells are mainly present in the TIGITlow CD8 T cells, indicating that belrestotug is mainly depleting the exhausted CD8 T cells (Fig. 1D).

**Figure 1:**
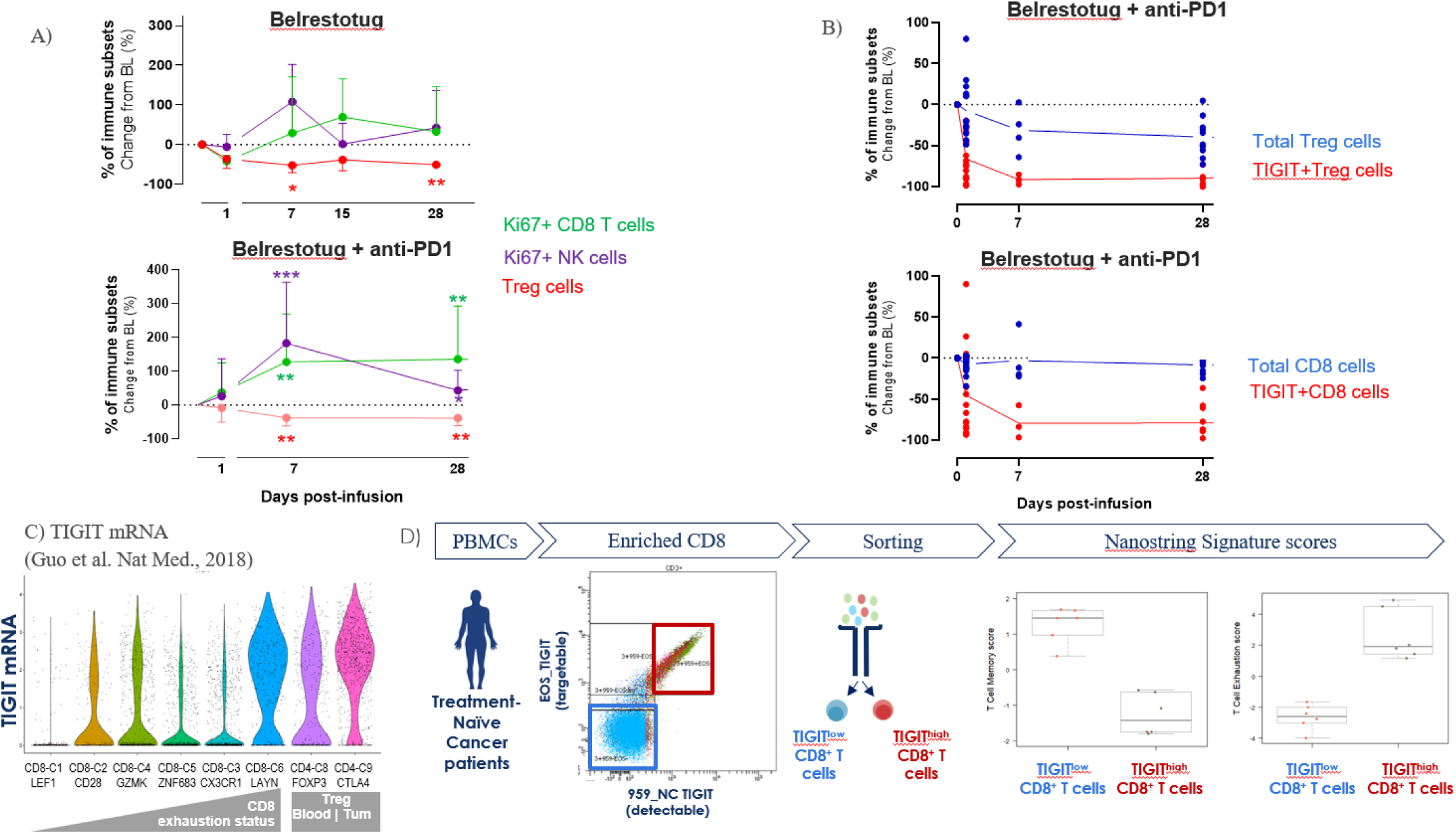
Effect on immune cell populations in the peripheral blood of patients receiving belrestotug monotherapy and in combination with anti-PD1. a. Belrestotug depletes Treg cells and induces T/NK cell proliferation in the periphery, both in monotherapy and in combination with anti-PD1 (paired T test * p<0,05, ** p<0,005, *** p<0,0001 ) b. Peripheral TIGIT+CD8T cells and Treg cells are downregulated upon combination treatment with Belrestotug and anti-PD1 c. Violin plot showing TIGIT mRNA levels from Guo et al^18^ cross different CD8 differentiation states and peripheral vs intratumoral Tregs. d. Most of the TIGIT^high^CD8 T cells that are targeted by Belrestotug have an exhausted phenotype

### Dual Blockade of TIGIT and PD-1 impacts Tregs and CD8 in patients’ tumors

The origin of intratumoral Treg cells is only partially understood. In breast cancer patients, intratumoral Tregs share similar features with peripheral Tregs, which supports their relationship ^21^. Thus, based on the changes observed in the peripheral blood, we next analyzed intratumoral Tregs within on-treatment biopsies using TIGIT and FoxP3 (Treg marker) staining immunohistochemistry (IHC). We observed that belrestotug monotherapy significantly decreases TIGIT+Treg density resulting in 2 to 4 fold reduction (Fig. 2A). Interestingly, a-PD-1 therapy elevates the number of Tregs in metastatic melanoma patients ^11^. Thus, we sought to evaluate how belrestotug could alter TIGIT+Tregs density when combined with a-PD-1. As shown in Fig. 2B, TIGIT+ Treg densities were significantly downmodulated by the combination treatment, which suggests that a-TIGIT can elicit its effect despite the potential Treg amplification effect exerted by a-PD-1. This downmodulation effect is more pronounced in the stromal region compared to the tumor nest, in agreement with recent data suggesting that tumor infiltrating Tregs prevalently distribute in the stroma in NSCLC ^12^. In contrast, CD8 T cell densities are elevated in both the stroma and tumor nest (Fig. 2C), likely as a result of the a-PD-1 effect.

**Figure 2:**
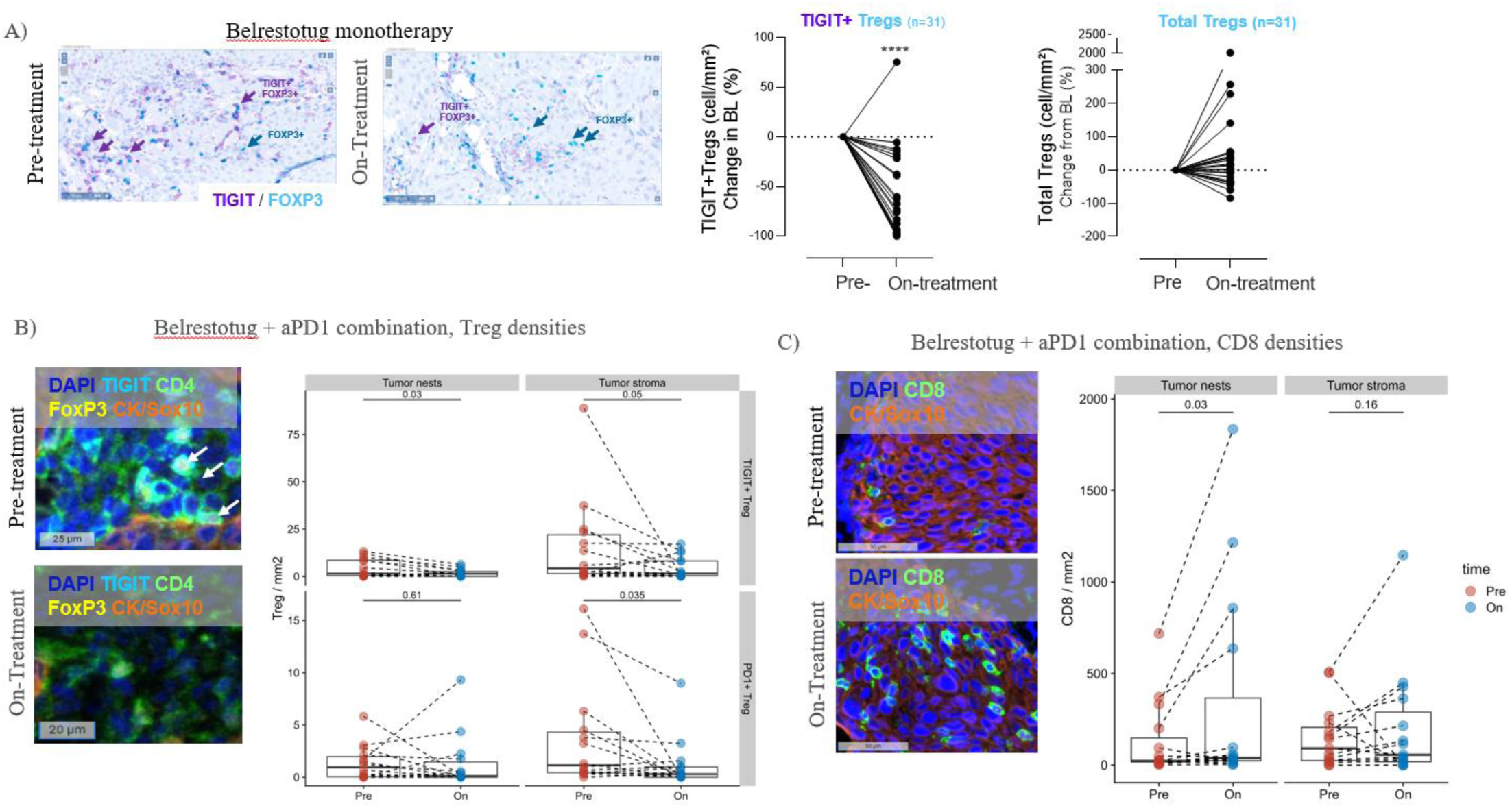
Belrestotug alone or in combination with aPD1 depletes immunosupressive Tregs especially in the stoma and combination with aPD1increases CD8 densities. Left, representative tissue images showing TIGIT and FoxP3 staining infiltration in the tumor before and after treatment. Right, percentage change from baseline of TIGIT+FoxP3+ cells/mm2 and total FoxP3+ from pre and on-treatment biopsies. Statistics: Paired T test. a. Left, representative images from 8-plex Immune fluorescence analyzed with Ultivue from patients treated with Belrestotug and anti-PD1 combination, multichannel image depicting DAPI, TIGIT, FoxP3, CK/SOX10 staining. Right, changes in cells density of TIGIT+Tregs and PD-1+Tregs between pre-treatment biopsies and paired on-treatment biopsies, and charts comparing densities at different annotated tumor areas (tumor nests and tumor stroma). Statistics: Mann-Whitney U Test. b. Left, representative images from 8-plex multi-fluorescence analysis with Ultivue from patients treated with Belrestotug and anti-PD1 combination, multichannel image depicting DAPI, CD8, CK/SOX10. Right, changes in cells density of TIGIT+Tregs and PD-1+Tregs between pre-treatment biopsies and paired on-treatment biopsies, and charts comparing densities at different annotated tumor areas (tumor nests and tumor stroma). Statistics: Mann-Whitney U Test

### Impact of combination therapy on Treg-CD8 T cell spatial localization and tumor reduction

In melanoma patients, Tregs are found to be close to CD8 T cell clusters ^11^. Thus, we further explored how the combination treatment could affect Treg-CD8 T cells spatial localization using multiplex immunofluorescence. We analyzed the percentage of CD8 T cells in the vicinity of Treg cells and observed that the treatment decreased the proportion of CD8 T-cells proximal to at least one Treg cell in a radius of 100µm for patients included in this analysis (p=0.022; Fig. 3A). We hypothesized that the pharmacodynamic relationship between CD8 T cells and Tregs could be assessed in baseline biopsies and tested whether their proximity correlated with tumor reduction. We calculated the distance (µM) between CD8 T-cells and Tregs in the stroma prior to treatment (Fig. 3B) and observed that closer CD8 T cells-Tregs distances were associated with tumor volume reduction. Moreover, CD8 T cells-Treg distances were inversely correlated with CD8 T-cells densities (Spearman correlation coefficient= −0.91, p=3.3e-07 ; Sup.Fig2) and higher CD8 T-cells densities were also associated with tumor reduction (p=9.6e-5) (Fig. 3B). Interestingly CD8 T-cells densities were strongly correlated with total Treg densities as quantified by IHC (Spearman correlation coefficient=0,83, p=4.4e-07, Sup.Fig.2) suggesting that CD8 T-cells densities could be a predictive biomarker for response to belrestotug plus a-PD-1 combination. Tumor infiltrating CD8 T-cells density and distribution allows to stratify tumors in 3 different immunotypes: inflamed, excluded and desert. Several studies have suggested that upon PD-(L)-1 blockade most responses occur in patients with inflamed tumor immunotype ^22^ ^23^. The excluded immunotype is also characterized by high T cell infiltration that is more restricted to the stromal area, where regulatory T-regs are also located. Thus, we reasoned that as the combination induced major changes in the stromal area, the patients with excluded immunotype may benefit from the combination therapy in addition to the inflamed immunotype. Consistent with this hypothesis, we observed that a higher frequency of patients with tumor reduction presented an inflamed and excluded immunotype (Fig. 3D) compared to desert patients. Interestingly, despite testing a late-stage population that is mostly PD-L1 low or null, we could observe a statistical clinical benefit as shown by increase median progression free survival in the inflamed and excluded patient population compared to the desert subgroup (Fig. 3D). These data suggest that belrestotug addition to a-PD-1 may be beneficial to the excluded immunotypes and potentially to the inflamed one in those PD-L1 low or null solid tumors.

**Figure 3:**
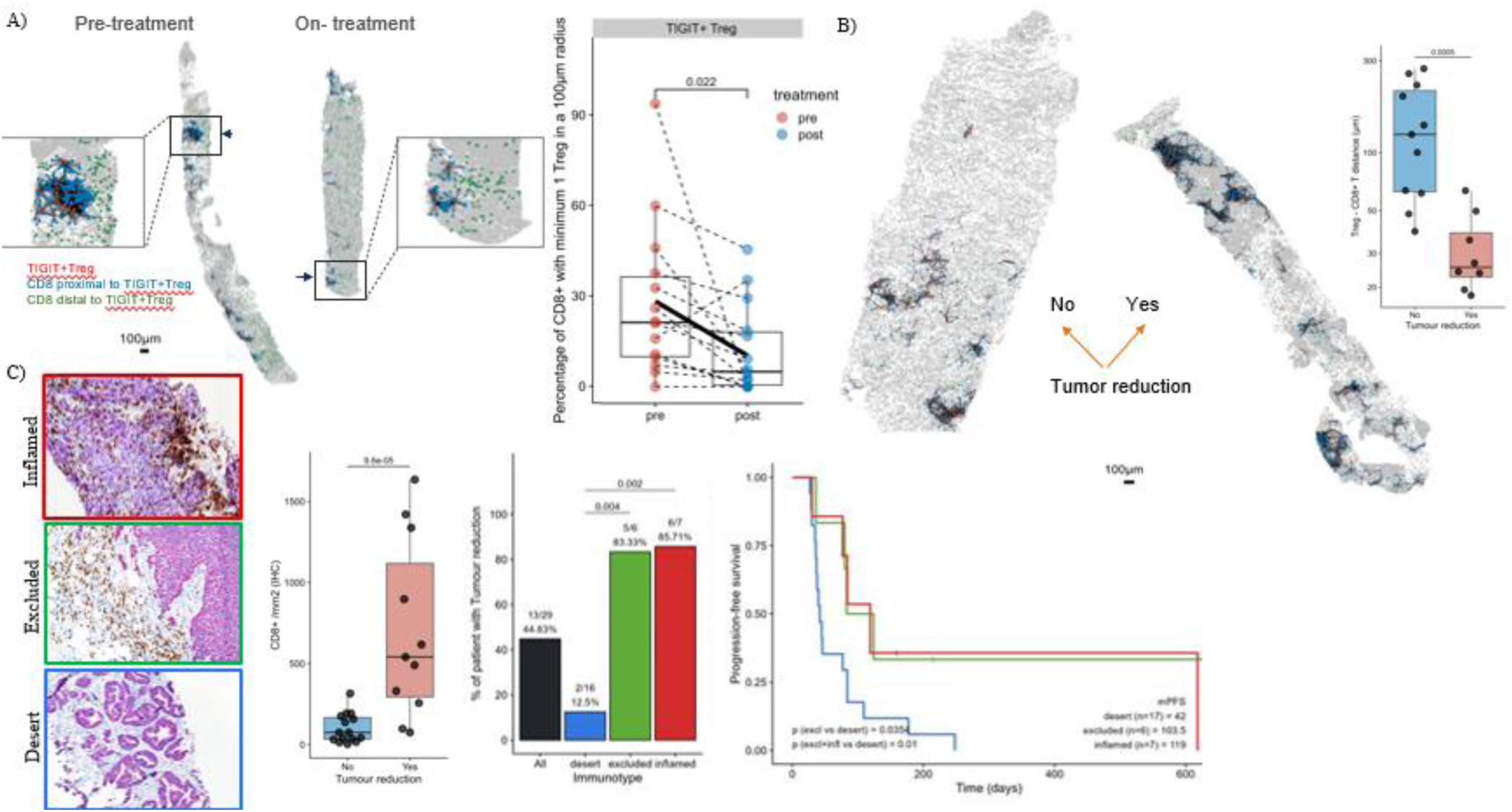
CD8 based Immunotypes associates with tumor reduction in belrestotug + aPD-1 treated patients. a. Representative coordinate map depicting the spatial distribution TIGIT+Tregs (red), CD8 proximal to TIGIT+Treg (radius <100um; blue), CD8 distal to TIGIT+Tregs (radius >100um; green). Quantification from paired biopsies of CD8+ cells frequency with a minimum of one Treg in a 100uM radius (statistics: linear mixed model). Dash lines connect paired biopsies, black lines represent the mean. b. Representative coordinate map depicting the spatial distribution and CD8-Treg distance in pre-treatment biopsies. Box plot showing the CD8-Treg distance in pre-treatment biopsies in patients stratified by tumor reduction by RECIST1.1. (statistics: linear mixed model) c. Representative images from CD8/PanCK stanning from different pre-treatment biopsies with inflamed, excluded and desert immunotypes from late-stage cancer patients. Box plot showing the CD8 density (cells/mm2) in pre-treatment biopsies in patients stratified by tumor reduction by RECIST1.1. Statistics: Mann-Whithney U. Bar graph showing frequency of immunotype distribution cases across patients with tumor reduction. Statistics: Fisher exact test. PFS analysis of the patient population stratified by immunotypes. Statistics: cox regression.

### a-TIGIT plus a-PD-1 combination differentially upregulate IFNγ signaling

We sought to understand the effect of the combination therapy on the CD8 T-cells activation status. We generated spatial transcriptomics data using the Visium platform (10X Genomics Inc.). Paired tumor biopsies from 9 patients treated with belrestotug plus a-PD-1 combination, across multiple cancer indications. Focusing specifically on the stroma, we observed a downmodulation of the Treg signature alongside an increase in signatures associated with CD8 T cells and TAMs. The significant increase in total CD8 T cell signature (Fig. 4B) further confirms the augmented CD8 T cells numbers observed by spatial proteomics (Fig 2C). There is also an upregulation in CD8 T-cells interferon gamma signature (Fig 4B), that supports an enrichment in activated CD8 T-cells^14^. The treatment significantly upregulated also the TAM signature^17^ in stromal spots (Fig 4B). Next, we performed a differential gene expression analysis between pre and on-treatment biopsies, as shown in the volcano plot on Fig. 4C. CCR5 axis genes, CCL5 and CCL4, were upregulated on-treatment. Recent research suggests that immunotherapy activated intratumoral CD8 T-cells recruit macrophages via CCR5 signaling and skew them towards a late-stage activated and tumoricidal phenotype ^24^. Thus, the differential gene expression analysis results suggest that the TAM activation may be dependent on the CD8 T-cells activation via the CCR5 axis, as the remodeling of the tumor micro-environment might be primarily initiated by the changes in the CD8 T-cells compartment. We applied 19 curated gene signatures derived from 14 CD8 T-cells clusters defined across 16 cancer indications from a recent single cell RNAseq atlas ^14^. The normalized score from the gene set enrichment analysis (GSEA) revealed a treatment mediated upregulation of several gene signatures for CD8 T-cells, including IFNγ response, stress response and activation/effector function (FDR < 0.05) (Fig 4D). a-PD-1 monotherapy markedly upregulates the stress response pathway ^24^ indicating that the upregulation of additional pathways and signatures could be the result of the combination with the a-TIGIT treatment.

**Figure 4:**
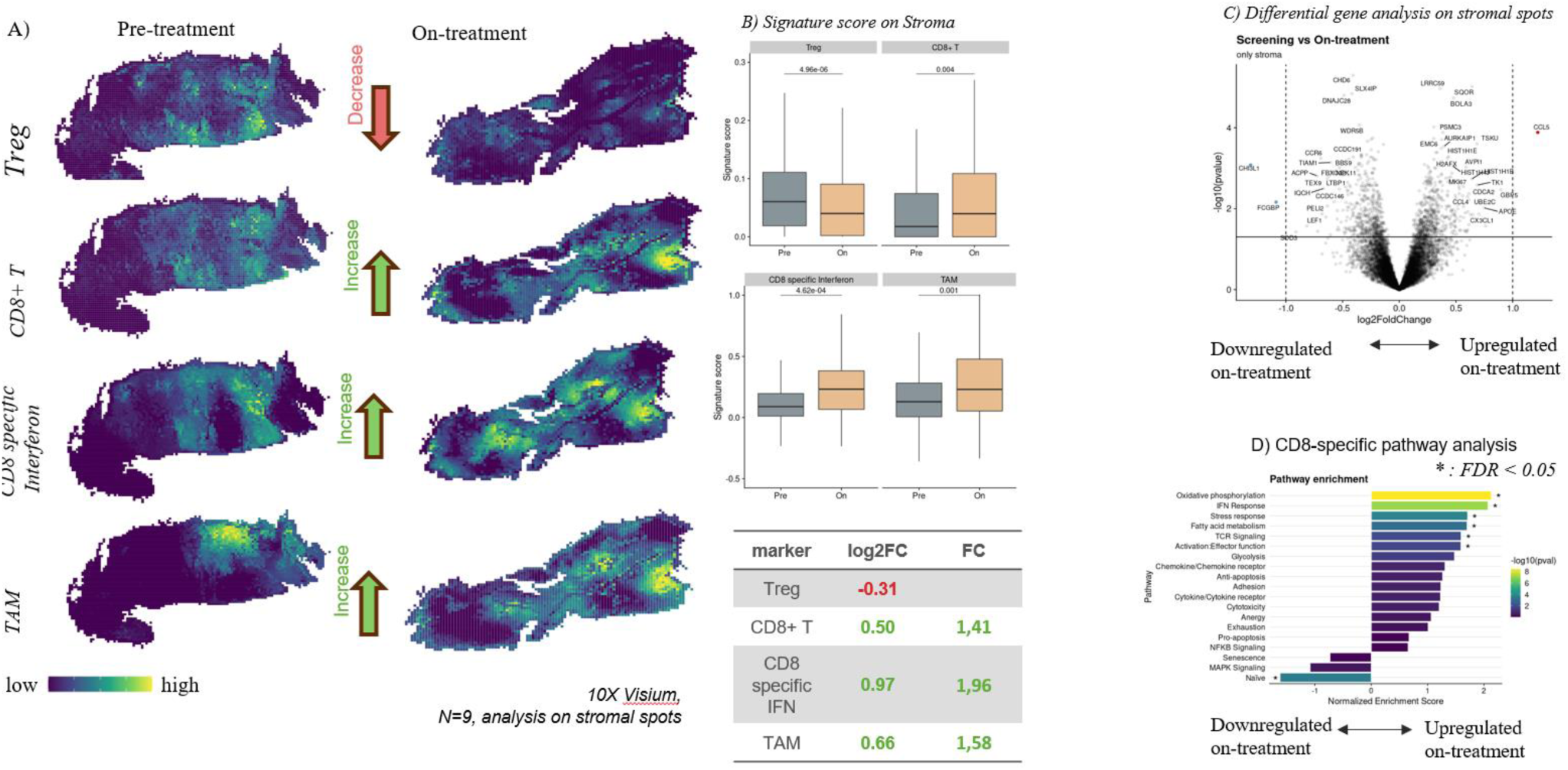
Spatial transcriptomics confirms Treg depletion and CD8+ increase in the tumor stroma and modifications of myeloid composition upon treatment. a. Representative tissue sections from pre-treatment biopsy (left) and on-treatment biopsy (right). Scores represent CD8, CD8 specific interferon ^14^ and TAM ^17^. b. Box plots representing treatment induced changes in Treg , CD8, CD8 specific interferon^14^ and TAM signatures ^17^ between pre-treatment and on-treatment biopsies in stromal spots. c. Volcano plot showing the Differential gene expression analysis induced by treatment on stromal spots d. Normalized enrichment score induced by combination treatment for CD8 specific pathways as defined in ^14^. N=9 paired pre- and on-treatment biopsies from 9 patients, from 6 different cancer indications). Statistical analysis were performed using LMM (linear effect mixed models as defined in Material and methods)

### a-TIGIT monotherapy induces phenotypic changes in intratumoral CD8 T-cells *in vivo*

To further elucidate the TIGIT blockade effect on intratumoral CD8 T-cells pathways, we analyzed CD8 T-cells subsets in murine CT26 tumors at different times after treatment with a-TIGIT mAb, comparing a-TIGIT Fc variants as monotherapy or in combination with a-PD-1. *In vivo* murine tumor models, reported that the dual TIGIT and PD-1 blockade can enhance IFNγ production in tumor-infiltrating CD8 T cells ^13^, in agreement with previous reports ^25^. At the first timepoint evaluated, anti-TIGIT Fc live treatment increased several CD8 T-cells subsets, including CD8 T naïve, T-stem and T-effector cells (Fig 5B), whose gating strategy is shown in Sup.Fig.3. The addition of a-PD-1 resulted in a similar effect over those cell subsets, which supports the role of a-TIGIT in modulating CD8 T-cells infiltration and activation. Interestingly, a-TIGIT effect differs across variants. Thus, the a-TIGIT Fc live variant, able to engage activatory Fc γ receptors, exerts a more pronounced increase of CD8 T naïve and Tstem than the mouse IgG1 or a-TIGIT Fc dead version. As those cell subsets do not express TIGIT, these findings might suggest a cell extrinsic effect that is independent of TIGIT blockade (Fig 5B). Concomitantly, after the first dosing, the exhausted CD8 T-cells subset co-expressing TIGIT+PD-1+TIM-3+ appeared downmodulated by a-TIGIT Fc-live variant. Thus a-TIGIT Fc-live might support CD8 T stem cell amplification, an effect also associated with the a-PD-1 blockade ^26^. The analysis after multiple dosing revealed a sustained increase of the T-stem population. Effector T-cells expressing PD-1 and TIM-3 were differently modulated, a-TIGIT Fc-live monotherapy triggered a sustained increase after 3 doses. This data is in line with other studies that suggest that the combination of a-TIGIT Fc live with a-PD-(L)-1 inhibitors shift tumor infiltrating CD8 T cells towards a memory-like state, driving them away from an exhaustion program ^17^.

**Figure 5:**
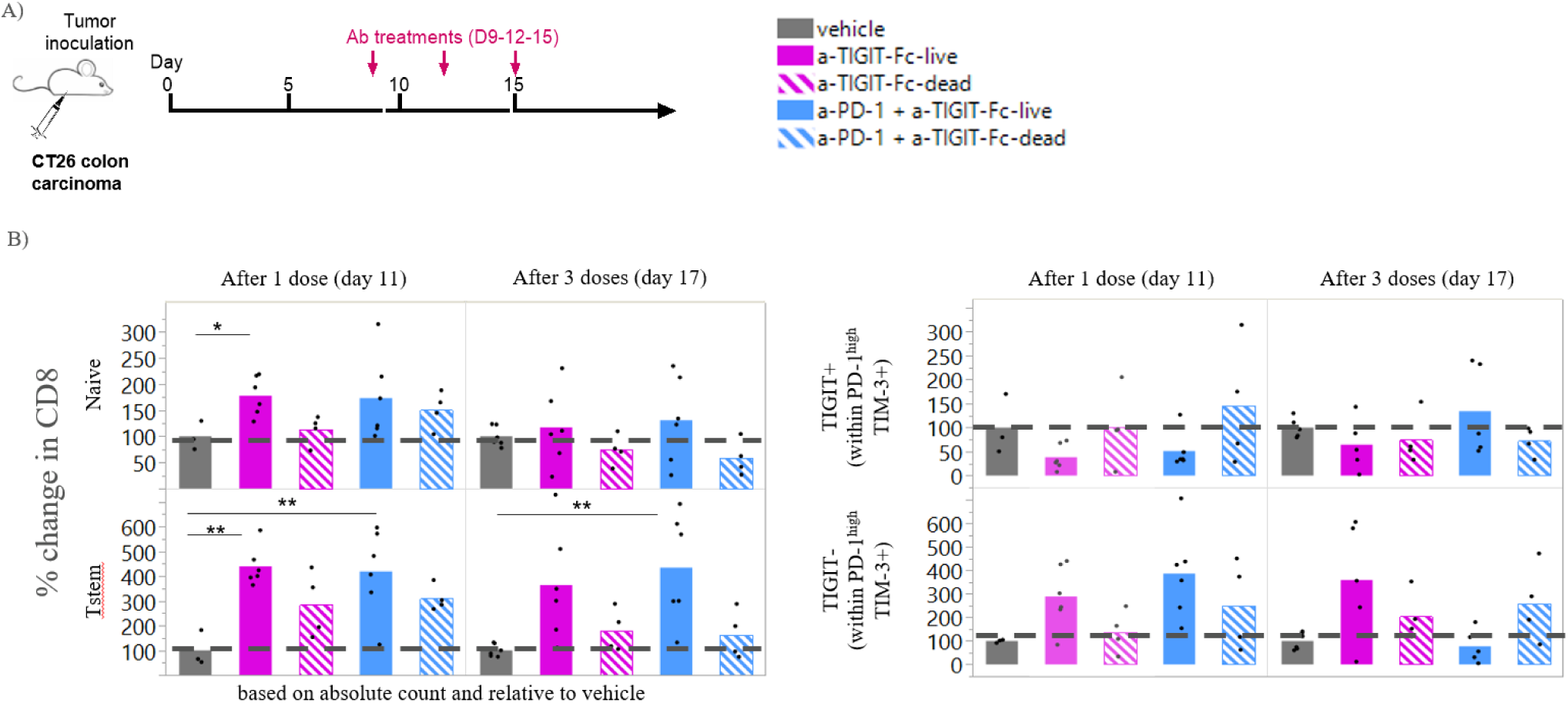
a-TIGIT + a-PD-1 treatment decreases CD8 exhausted cells while increasing Tstem population in TME. a. Experimental setup b. Phenotypic analysis of infiltrating CD8 cell subsets in murine CT26 tumors at different timepoints after treatment, by flow cytometry (n=4-6 mice per treatment group and timepoint). One-way Anova * p<0,05, ** p<0,005.

## Discussion

During the past years multiple TIGIT-targeting antibodies entered clinical development across different cancer indications, alone or in combination with PD(L)-L1 inhibitors, have demonstrated potential in augmenting immune response in cancer malignancies ^27^ .

In 2 clinical studies (NCT04335253, NCT05060432), we analyzed fresh blood and paired biopsy samples originating from late-stage all solid tumors cancer patients (mostly PD-L1 low or negative) and observed that belrestotug alone or in combination with a-PD-1 can deplete peripheral Tregs. Its effect of inducing proliferating T and NK cells in blood is further enhanced in combination. Intratumorally, belrestotug reduces TIGIT+Tregs in monotherapy. When belrestotug is combined with a-PD-1, there is a reduction in Treg cells, preferentially localized in the stroma, and proximal to CD8 T cells. Since Treg and CD8 T-cell densities correlate within tumors, CD8 T-cell density and distribution may serve as proxies for the frequency of Tregs near CD8 T-cells. Our data analysis revealed that the inflamed and excluded immunotypes may be predictive biomarkers of belrestotug plus a-PD-1 combination, as they are characterized by elevated numbers of CD8 T cells in the stromal region. Mechanistically, the combination treatment remodeled the tumor microenvironment. Stromal Tregs reduction was accompanied by CD8 T-cells IFNγ and TAM signatures increase. The transcriptomic analysis identified members of CCR5 axis upregulated by the treatment. Activation of CD8 T-cells leads to upregulation of the CCR5 axis, which in turn facilitates further tumor remodeling through the recruitment and activation of macrophages^24^. Interestingly, in advanced cancer patients the treatment augmented different CD8 T-cells pathways in the stroma regions, including IFNγ response, stress response and activation/effector functions. Intratumorally, there are several divergent differentiation CD8 trajectories starting from the CD8 T naïve compartment. Those differentiation pathways result in CD8 exhausted, CD8 effector and CD8 stress response T-cells. a-PD-1 monotherapy markedly induces the CD8 T-cells stress response signature ^14^, while the combination with belrestotug also augments CD8 IFNγ response and the CD8 activation/effector. These findings suggest that the combination might alter the proportions of differentiated CD8 T-cells across those divergent pathways. In mouse models, a-TIGIT monotherapy induces an effector-like CD8 T-cells subset after a single dosing and to similar levels than in combination with the a-PD-1 treatment, while reducing the CD8 exhausted T-cells compartment.Although this study’s patient population differs from those in major trials and lacks long-term clinical data, it demonstrates that combined TIGIT and PD1 inhibition can remodel the TME and may benefit certain patient subgroups. Further investigation in larger patient cohorts would help support the biomarker hypothesis for belrestotug combination with aPD1.

## Methods

### Clinical trials, patient cohort and response assessment

Tissue and blood samples were obtained from patients enrolled in belrestotug monotherapy study (NCT04335253) or in combination study (NCT05060432). Clinical studies were performed in accordance with the Declaration of Helsinki and Good Clinical Practice Guidelines. Patients in belrestotug monotherapy study received escalating doses of belrestotug alone every 3 or every 4 weeks. Patients in combination study received escalating doses of belrestotug in combination with standard of care 200 mg Pembrolizumab or 500 mg Dostarlimab, every 3 weeks. Response was evaluated as per RECIST v1.1 for solid tumors.

### Flow cytometry in whole blood

Up to 16 patients were included in the analysis, all of them received the same dose of belrestotug either alone or in combination with a-PD-1 (either pembrolizumab or dostarlimab). Samples were collected before treatment and at different times after treatment 24h, 7 days, 15days and 28 days. Each sample was analyzed with different custom panels as described in Sup. table 1. 200ul of Whole blood were stained with its corresponding antibody cocktail, for 30min at room temperature, then incubated with Pharmalyse buffer for 15min at room temperature, centrifuged washed and resuspended on Stain buffer prior to acquisition on FACSCanto. For the analysis of immune activation, after staining with surface markers, samples were fixed and permeabilized using the FoxP3 transcription factor fixation kit according to manufacturer’s instructions, and stained for intracellular marker for 30 min at room temperature. Then samples were centrifuged, washed and resuspended in stain buffer for acquisition in FACS Aurora.

### CD8 sorting and transcriptomics

Peripheral blood mononuclear cells (PBMC) were prepared from treatment-naïve melanoma (n=2) and head and neck (n=4) patients. CD8 T cell were enriched with StemCell T cell isolation kit (17951, StemCell) according to the manufacturer’s instructions. CD8 T cells were stained for 30 min at 4°C (with CD8, clone SK1, and TIGIT clones EOS88448, and ADI-32959), washed and resuspend at 10*10^6^cells/ml for sorting. a FACS Aria III cell sorter (BD Biosciences) based on the level of TIGIT Sup.Fi 1.

Sorted cells were resuspended in RLT buffer and RNA was extracted with RNeasy plus micro kit from Qiagen (74034), and analyzed by Nanostring with the nCounter immune Exhaustion panel.

### Duplex IHC TIGIT/FoxP3

Biopsies from up to 31 patients enrolled in Phase I study IO-002 (NCT04335253) were collected and processed at pre-treatment and at C2D3. 4-µm paraffin slides were stained using a Ventana Discovery Ultra platform (Ventana, Roche Diagnostics). After antigen retrieval, slides were incubated with FoxP3 antibody (SP97; Abcam) and TIGIT antibody (E5Y1W; Cell Signalling Technology). FoxP3 and TIGIT expressing cells were quantified with an automated scoring application (APP) from Visiopharm software performed by an Imaging scientist under supervision of a pathologist.

### 8-plex Immunofluorescence stating and image analysis

Biopsies from up to 19 patients enrolled in Phase II study TIG-006 (NCT05060432) were processed at pre-treatment and at C2D3. FFPE tissue sections were stained with an off-the shelf Ultivue panel including Ki67, CD4, FoxP3, HLA-DR, CD8, PD-1, and CK/Sox10, applying its patented InSituPlex^®^ technology^28^. TIGIT detection was included using the commercially available clone (E5Y1W, Cell signaling).Cell detection and classification were performed using Ultivue’s deep learning models.

### Spatial analysis of multiplex immunofluorescence

Spatial coordinates of CD8+ T cells and Tregs were extracted, and the Euclidean distance was calculated between each Treg cell and its nearest CD8+ T cell. To evaluate the relationship between spatial immune cell organization and tumor reduction, a linear mixed-effects model was applied with the formula: distance ∼ tumor reduction_status + (1|patient)

In this model, distance represents the nearest-neighbor CD8–Treg distance, ∼ tumor reduction_status denotes binary tumor reduction status, and patient was included as a random effect to account for inter-patient variability. A p-value was calculated to test for significant differences in CD8–Treg spatial proximity between tumor reduction and non-reduction groups. This model was implemented using the lmerTest R package.

### CD8 IHC and Immunotype

Biopsies from up to 29 patients enrolled in Phase II study TIG-006 (NCT05060432) were processed at pre-treatment. FFPE tissue sections were stained for CD8 and pan-cytokeratin (panCK) using a CD8-panCK duplex chromogenic IHC assay developed and validated by CellCarta (Wilrijk, Belgium). Staining was performed on the Ventana Discovery Ultra platform (Roche Diagnostics). The CD8 density analysis was performed using an analysis protocol package (APP) in Visiopharm. The CD8 immunotype or immunophenotype^22^ is a pathologist-based scoring system that classifies samples into “desert”, “excluded” or “inflamed” groups. The method was developed and validated by CellCarta and it has previously described ^29^.

### RNA Quality Assessment and sequencing for Visium analysis

Biopsies from up to 9 patients enrolled in Phase II study TIG-006 (NCT05060432) were processed at pre-treatment and C2D3. Up to five FFPE sections per tissue sample were used, for H&E staining and RNA extraction. For each sample a capture area of 6.5 x 6.5 mm (5.000 spots) was annotated by a pathologist. RNA extraction was performed with the High Pure FFPET RNA extraction Kit (Roche). DV200 values were evaluated using BioAnalyzer (Agilent Technologies), and only samples with DV200 values superior to 30% were included in the analysis. After library preparation, the generated libraries were sequenced on the Illumina NovaSeq 6000 platform.

### Spatial Transcriptomics analysis

Spatially resolved transcriptome information from 10X Genomics Visium platform was processed using Space Ranger software (v2.1.1, 10X Genomics). Raw sequencing reads were mapped to the human reference transcriptome (GRCh38; refdata-gex-GRCh38-2020-A) using 10X spaceranger count pipeline. Tumor and stroma regions were manually annotated by an experienced pathologist and used to relate gene expression patterns to tissue architecture. Downstream analysis was performed using the Seurat R package (v5.1.0). Spots were filtered to retain high-quality data based on the following criteria: total counts of detected transcripts (nCount_Spatial) > 5,000, number of detected genes (nFeature_Spatial) > 500, and percentage of mitochondrial gene expression (percent.mt) < 20%. Normalization of the data was performed using Seurat’s NormalizeData() function with its default settings. Variable features were identified using FindVariableFeatures() with “vst” as the selection strategy and retaining the top 2,000 genes for subsequent analysis. The data were then scaled using Seurat’s ScaleData() function. To integrate data across samples and conditions, individual Seurat objects were aligned using canonical correlation analysis (CCA). Integration anchors were identified with FindIntegrationAnchors() (reduction = “cca”, dims = 1:30), and datasets were merged using IntegrateData() with k.weight = 50 to balance sample-specific and shared variation. The integrated dataset was used for downstream analysis.Gene signature scores were computed for each spatial spot using Seurat’s AddModuleScore() function. To assess treatment-associated changes in gene expression and gene signature expression, we performed a differential analysis of module scores between pre- and post-treatment samples. A linear mixed-effects model was applied using the following formula: expression_*score ∼ time (pre vs post) + (1|patient_id) ;* where expression refers to gene expression and score refers to the module score for each spot, time represents the treatment condition (pre- or post-treatment), and patient_id was modeled as a random intercept to account for repeated measures from the same individual. This model was implemented using the lmerTest R package.

### Tumor growth in *in vivo* models and characterization of immune infiltrate

Experiments with in vivo models were performed as described previously ^13^. Briefly, eight-week-old female BALB/c (Charles River Laboratories) were inoculated subcutaneously with 5×105 CT26 (ATCC CRL-2638TM ) tumor cells. Tumor volume was measured three times per week. Mice were randomized when tumors were established and treated using PBS or mIgG2a isotype (200 mg/mouse, Bio X Cell BE0085), anti-TIGIT mIgG2a (20 or 200 mg/mouse), anti-TIGIT mIgG1 (200 mg/mouse), anti-TIGIT hIgG1 (200 mg/mouse), anti-TIGIT hIgG1-N297A (200 mg/mouse), anti–PD-1 (200 mg/mouse, Bio X Cell BE0146). All antibodies were diluted in PBS prior intraperitoneal injection every three days for a total of three injections. Two days after first and third antibody treatments (days 11 and 17), 4-6 CT26 tumor-bearing micewere sacrificed and tumors were processed for staining using the antibody cocktails described in sup. table 2.

## Data Availability

All data produced in the present study are available upon reasonable request to the authors

## Supplementary Figures

**Supplementary Figure 1:**
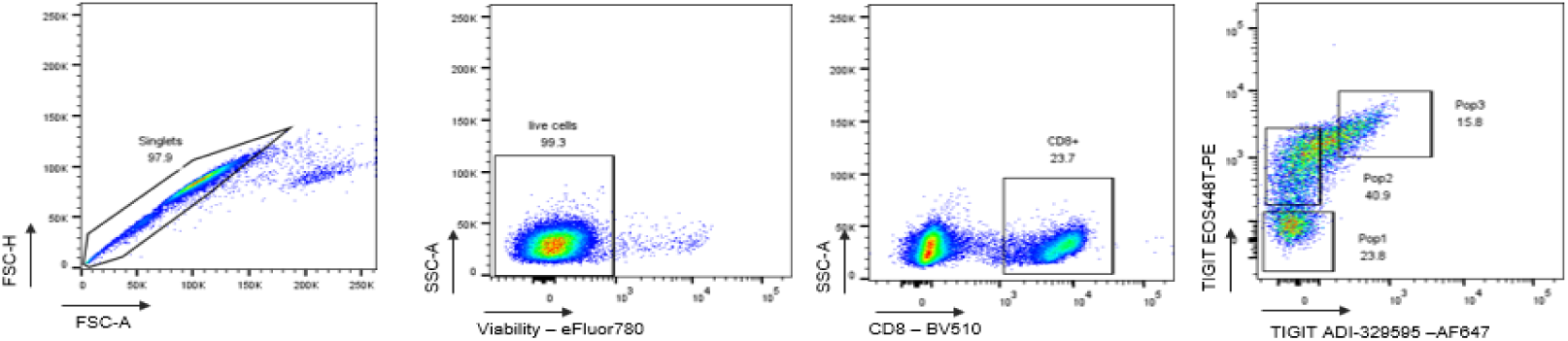
gating strategy sorting TIGIT populations on CD8+ cells

**Supplementary Figure 2:**
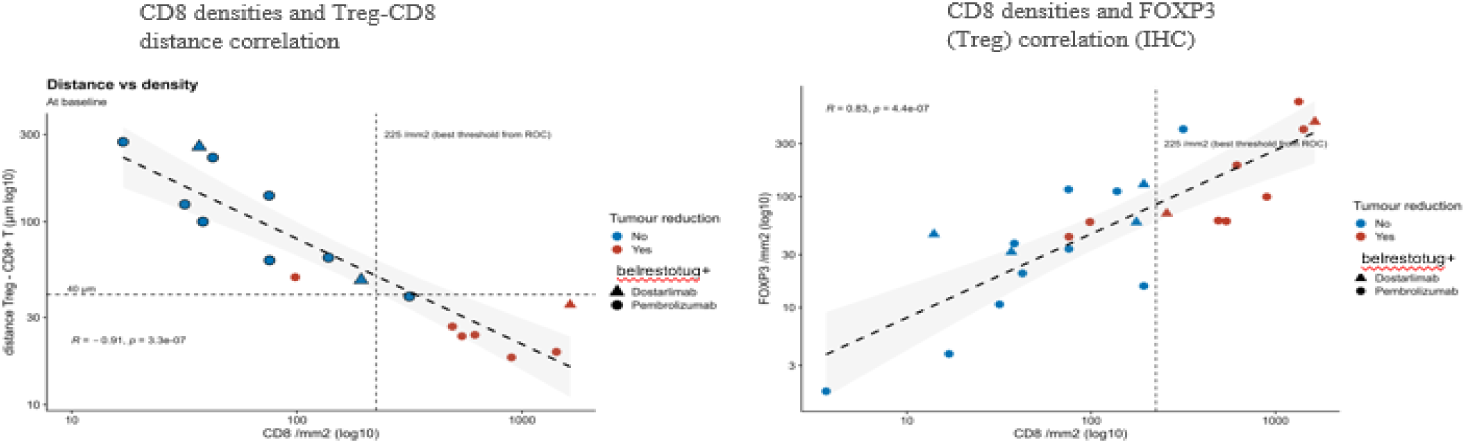
Multiplex showing belrestotug in combination with anti-PD1 decreases the frequency of CD8 T cells proximal to TIGIT+ Tegs in tumor stroma

**Supplementary Figure 3:**
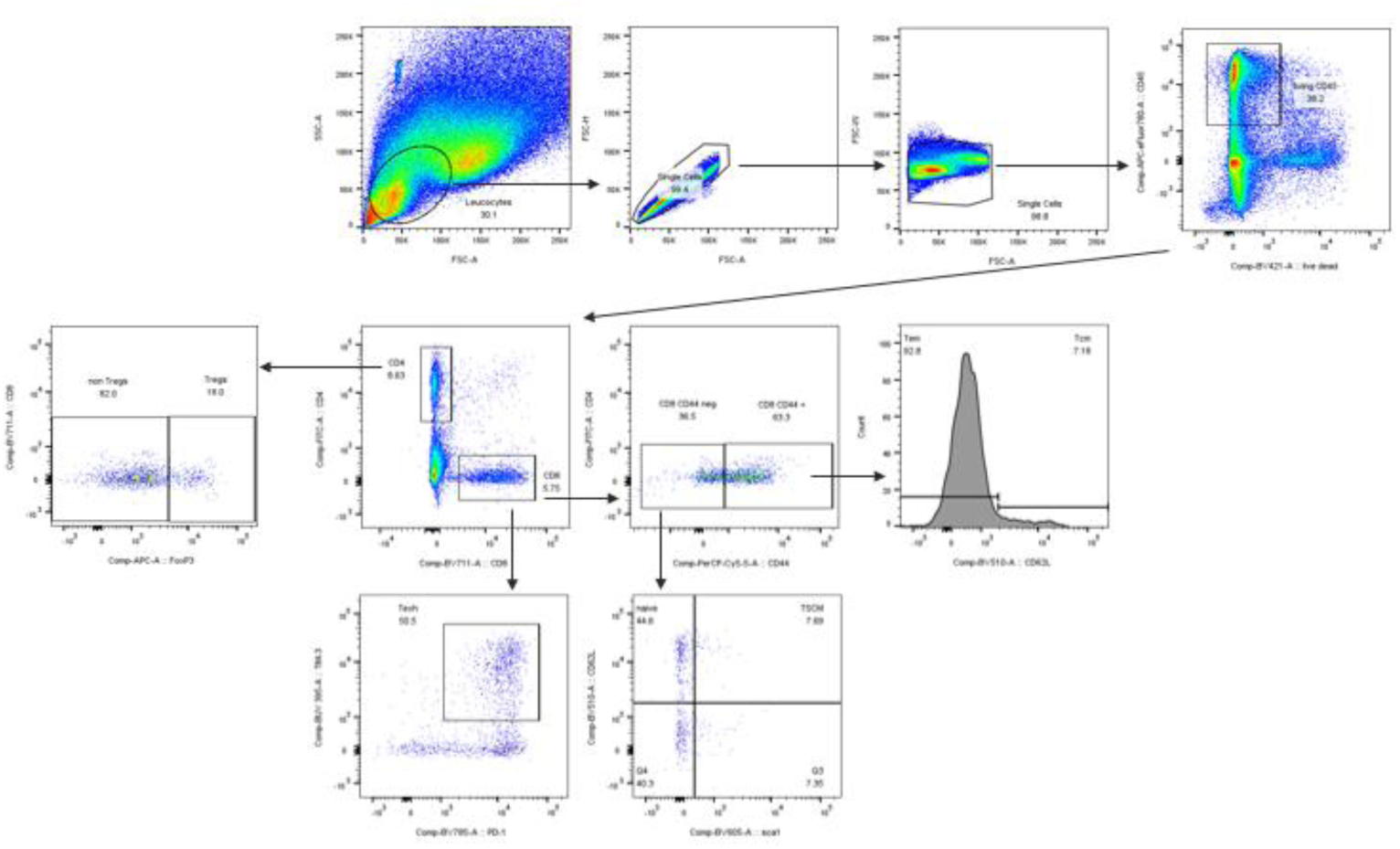
Gating strategy *in vivo* Tregs and CD8

**Supplementary Table 1:**
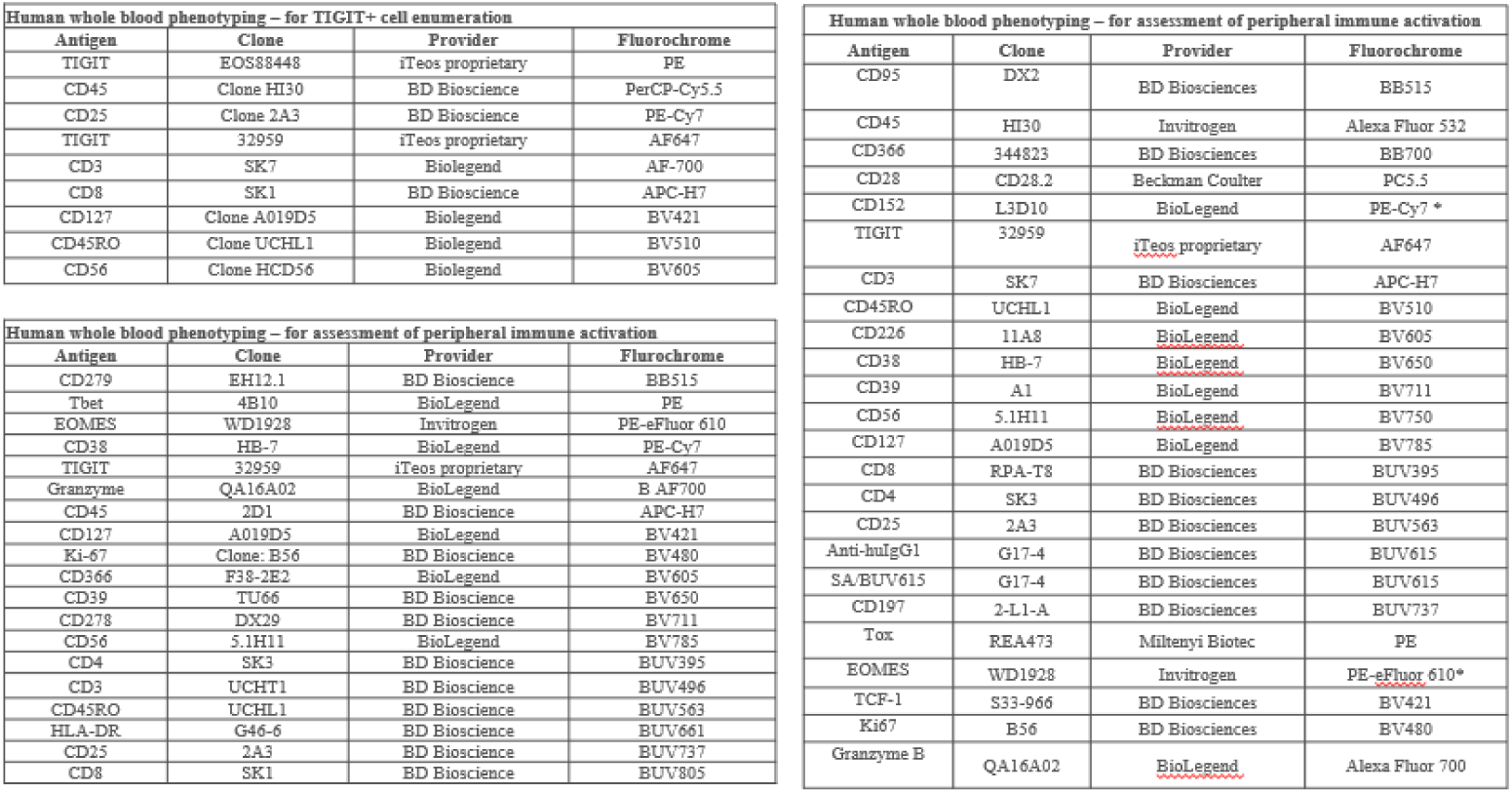
phenotyping of whole blood samples from cancer patients treated with belrestotug monotherapy or combination with a-PD-1.

**Supplementary Table 2:**
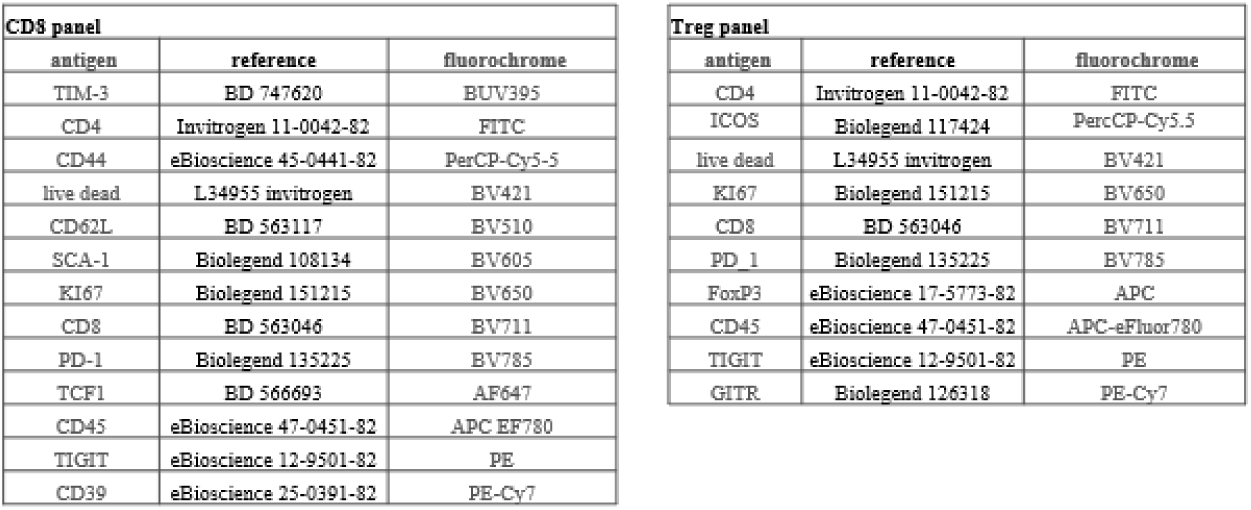
for immune infiltrate in *in vivo* models (related to figure 5)

## References

1. Shergold, A. L., Millar, R. & Nibbs, R. J. B. Understanding and overcoming the resistance of cancer to PD-1/PD-L1 blockade. Pharmacol. Res. 145, 104258 (2019).

2. Yu, X. et al. The surface protein TIGIT suppresses T cell activation by promoting the generation of mature immunoregulatory dendritic cells. Nat Immunol 10, 48–57 (2009).

3. Banta, K. L. et al. Mechanistic convergence of the TIGIT and PD-1 inhibitory pathways necessitates co-blockade to optimize anti-tumor CD8+ T cell responses. Immunity 55, 512–526.e9 (2022).

4. Joller, N. et al. Treg cells expressing the coinhibitory molecule TIGIT selectively inhibit proinflammatory Th1 and Th17 cell responses. Immunity 40, 569–81 (2013).

5. Johnston, R. J. et al. The Immunoreceptor TIGIT Regulates Antitumor and Antiviral CD8+ T Cell Effector Function. Cancer Cell 26, 923–937 (2014).

6. Lucca, L. E., et al. TIGIT signaling restores suppressor function of Th1 Tregs. Jci Insight 4, e124427 (2019).

7. Fuhrman, C. A. et al. Divergent Phenotypes of Human Regulatory T Cells Expressing the Receptors TIGIT and CD226. J Immunol 195, 145–155 (2015).

8. Fourcade, J., et al. CD226 opposes TIGIT to disrupt Tregs in melanoma. Jci Insight 3, e121157 (2018).

9. Sade-Feldman, M. et al. Defining T Cell States Associated with Response to Checkpoint Immunotherapy in Melanoma. Cell 175, 998–1013.e20 (2018).

10. Kamada, T. et al. PD-1+ regulatory T cells amplified by PD-1 blockade promote hyperprogression of cancer. Proc. Natl. Acad. Sci. 116, 9999–10008 (2019).

11. Geels, S. N. et al. Interruption of the intratumor CD8+ T cell:Treg crosstalk improves the efficacy of PD-1 immunotherapy. Cancer Cell 42, 1051–1066.e7 (2024).

12. Devi-Marulkar, P. et al. Regulatory T cells infiltrate the tumor-induced tertiary lymphoïd structures and are associated with poor clinical outcome in NSCLC. Commun Biology 5, 1416 (2022).

13. Preillon, J. et al. Restoration of T-cell Effector Function, Depletion of Tregs, and Direct Killing of Tumor Cells: The Multiple Mechanisms of Action of a-TIGIT Antagonist Antibodies. Mol Cancer Ther 20, 121–131 (2021).

14. Chu, Y. et al. Pan-cancer T cell atlas links a cellular stress response state to immunotherapy resistance. Nat Med 1–13 (2023) doi:10.1038/s41591-023-02371-y.

15. Huang, A. C. et al. T-cell invigoration to tumour burden ratio associated with anti-PD-1 response. Nature 545, 60–65 (2017).

16. Kamphorst, A. O. et al. Proliferation of PD-1+ CD8 T cells in peripheral blood after PD-1– targeted therapy in lung cancer patients. Proc National Acad Sci 114, 4993–4998 (2017).

17. Guan, X. et al. Anti-TIGIT antibody improves PD-L1 blockade through myeloid and Treg cells. Nature 1–10 (2024) doi:10.1038/s41586-024-07121-9.

18. Guo, X. et al. Global characterization of T cells in non-small-cell lung cancer by single-cell sequencing. Nat Med 24, 978–985 (2018).

19. Beltra, J.-C. et al. Developmental Relationships of Four Exhausted CD8+ T Cell Subsets Reveals Underlying Transcriptional and Epigenetic Landscape Control Mechanisms. Immunity 52, 825–841.e8 (2020).

20. Galletti, G. et al. Two subsets of stem-like CD8+ memory T cell progenitors with distinct fate commitments in humans. Nat Immunol 1–11 (2020) doi:10.1038/s41590-020-0791-5.

21. Wang, L. et al. Connecting blood and intratumoral Treg cell activity in predicting future relapse in breast cancer. Nat Immunol 20, 1220–1230 (2019).

22. Mellman, I., Chen, D. S., Powles, T. & Turley, S. J. The cancer-immunity cycle: Indication, genotype, and immunotype. Immunity 56, 2188–2205 (2023).

23. Park, S. et al. Artificial Intelligence–Powered Spatial Analysis of Tumor-Infiltrating Lymphocytes as Complementary Biomarker for Immune Checkpoint Inhibition in Non–Small-Cell Lung Cancer. J. Clin. Oncol. 40, 1916–1928 (2022).

24. Elsas, M. J. van et al. Immunotherapy-activated T cells recruit and skew late-stage activated M1-like macrophages that are critical for therapeutic efficacy. Cancer Cell 42, 1032–1050.e10 (2024).

25. Johnston, R. J. et al. The Immunoreceptor TIGIT Regulates Antitumor and Antiviral CD8+ T Cell Effector Function. Cancer Cell 26, 923–937 (2014).

26. Sade-Feldman, M. et al. Defining T Cell States Associated with Response to Checkpoint Immunotherapy in Melanoma. Cell 175, 998–1013.e20 (2018).

27. Srikanth, G. et al. Promising New Anti-TIGIT Agents: Stealthy Allies in Cancer Immunotherapy. Clin. Transl. Sci. 18, e70212 (2025).

28. Manesse, M., Patel, K. K., Bobrow, M. & Downing, S. R. Biomarkers for Immunotherapy of Cancer, Methods and Protocols. Methods Mol. Biol. 2055, 585–592 (2019).

29. Li, X. et al. Automated tumor immunophenotyping predicts clinical benefit from anti-PD-L1 immunotherapy. J. Pathol. (2024) doi:10.1002/path.6274.

